# Childhood Trauma and *APOEε4* are Associated with Adolescent Brain Function, Posttraumatic Stress, and Alcohol-related Outcomes

**DOI:** 10.1101/2025.05.02.25326879

**Authors:** Zoe E. Neale, Megan E. Cooke, Jasmine Cárcamo, Maissa Trabilsy, Peter B. Barr, Chris Chatzinakos, David B. Chorlian, Weipeng Kuang, Gayathri Pandey, Alison M. Goate, Bernice Porjesz, Ananda B. Amstadter, Jacquelyn L. Meyers

## Abstract

Childhood trauma affects neurodevelopment and lifelong risk for posttraumatic stress disorder (PTSD) and alcohol use disorder (AUD). Changes in brain structures and function are observed in young carriers of *APOEε4*, the genetic factor most associated with Alzheimer’s disease. Longitudinal studies of *APOEε4*, childhood trauma, and neural connectivity in adolescence have not been explored. We studied 837 trauma-exposed participants (53% female) from the Collaborative Study on the Genetics of Alcoholism prospective sample, using latent growth curve models to assess associations of childhood trauma and *APOEε4* on repeated measures of frontal alpha EEG coherence (EEGc) throughout adolescence and young adulthood. Young adult AUD and PTSD symptoms were also examined. Results indicate childhood trauma and *APOEε4* are linked to neural connectivity, with effects differing by sex and trauma type. In females, sexual trauma was associated with a higher EEGc baseline but less growth, while *APOEε4* associated with lower right frontocentral (RFC) EEGc baseline and higher slope. In males, physical assault was associated with lower left frontocentral (LFC) EEGc baseline but increased growth, and non-assaultive trauma was linked to a lower RFC baseline and no association with growth. *APOEε4* was associated with lower LFC baseline and higher slope in males. Links between EEGc and AUD and PTSD were observed in both sexes, though effects differed in direction and strength. No significant trauma-by-*APOEε4* interactions emerged, nor direct links between *APOEε4* and PTSD or AUD. Findings highlight how EEGc may help explain connections between genetics, trauma, and psychopathology, guiding at-risk group identification and informing prevention strategies.

## Introduction

Childhood trauma, which is defined as a traumatic event (e.g., sexual assault, disaster exposure) occurring before the age of 18, has staggering consequences throughout the lifespan (Dye, 2018). Specifically, childhood trauma has long-term effects on neurodevelopment, leading to an increased risk of developing psychiatric and substance use disorders such as posttraumatic stress disorder (PTSD) and alcohol use disorder (AUD) (Bremner, 2006; Malarbi, Abu-Rayya, Muscara, & Stargatt, 2017; Meyers et al., 2018). Childhood exposure to traumatic events is linked to both structural and functional changes in the brain (Bremner, 2006; Cai et al., 2023; Carrion & Wong, 2012). Structurally, trauma is associated with decreased hippocampal volume, which is linked to memory loss and stress regulation (Teicher et al., 2003; Vythilingam et al., 2002). Functionally, brain changes include the overactivation of the amygdala and a blunted response in both the hippocampus and the medial frontal cortex (Bryant et al., 2008; Hayes, Hayes, & Mikedis, 2012; Shin et al., 2005). These brain regions are critical for stress response and emotional regulation. Additionally, PTSD is associated with increased risk of AUD, which may exacerbate long-term mental health challenges and further impact brain function (Brady, Back, & Coffey, 2004; Ouimette, Read, Wade, & Tirone, 2010). Chronic alcohol use has adverse effects on brain structures and a broad range of functions, especially those related to memory, executive functions, emotional and psychosocial skills, visuospatial cognition, and psychomotor abilities (Oscar-Berman & Marinković, 2007). Alcohol use is a top modifiable risk factor for dementia risk, emphasizing its critical importance in cognitive health across the lifespan (Schwarzinger et al., 2018). Therefore, advancing our knowledge on the complex and lasting impact of childhood trauma on brain structure and function could have far reaching impacts on mental health.

Given the profound effect of traumatic stress on aspects of brain functioning, it is unsurprising that PTSD is associated with an approximately 2-fold increase in risk for dementia (Günak et al., 2020). The impact of PTSD on risk for dementia may vary based on genetic risk, such that the effect of PTSD on dementia risk is more robust in carriers of Apolipoprotein isoform 4, commonly referred to as *APOEε4* (Averill et al., 2019; Lawrence, Rippey, Welikson, Pietrzak, & Adams, 2023; Logue et al., 2022). *APOEε4* is the most prominent genetic risk factor for the development of Alzheimer’s disease (AD) (Bellenguez et al., 2022; Corder et al., 1993; Lendon, Ashall, & Goate, 1997). Among individuals without AD, *APOEε4* is also associated with poorer cognition and complex health conditions (Reinvang, Winjevoll, Rootwelt, & Espeseth, 2010). While there is substantial evidence in older individuals, some studies have also demonstrated early developmental changes in brain structures and function that varied by *APOEε4* status (Iacono & Feltis, 2019), with healthy young ε*4* carriers (age 3-20) displaying smaller hippocampi, larger medial orbitofrontal cortical areas, and lower scores on executive function, working memory, and attention tasks (Chang et al., 2016). These neurobiological features associated with young carriers of *APOEε4* may contribute to an increased vulnerability to PTSD, as well as to AUD, where the interaction between genetics and environmental stressors can lead to maladaptive coping mechanisms involving heavy alcohol use (Averill et al., 2019; Bremner, 2006). Most studies investigating the impact of PTSD on the relationship between *APOEε4*, and neurocognitive outcomes have focused on older adults with less attention paid to the way trauma exposure and *APOEε4* may contribute to differences in cognitive functioning observed in adolescence and young adulthood. Accordingly, while we know that differences in cognition related to PTSD, alcohol use, and *APOEε4* have been detected in adolescence and young adulthood, we do not yet know the degree to which these influential risk factors may converge to influence brain functioning during this critical developmental period jointly. Therefore, there is a need to better understand how trauma, genetic risk, neurodevelopment, and psychiatric outcomes interact, with the hope of improving long-term outcomes in individuals affected by childhood trauma.

Many cognitive phenotypes have been employed to study AD and related dementias (ADRD). Electroencephalography (EEG) is a cost-effective and widely available method that provides functional data with a high temporal resolution which makes it an ideal modality to investigate complex diseases such as ADRD, PTSD, and AUD (Smit et al., 2021). EEG coherence (EEGc) is a heritable measure of functional connectivity derived by comparing the synchrony of EEG oscillations recorded at two different brain regions (Chorlian et al., 2007; Markovska-Simoska, Pop-Jordanova, & Pop-Jordanov, 2018). Previous studies have noted increased functional connectivity in cross-sectional studies of childhood trauma, AUD, and PTSD (Cook, Ciorciari, Varker, & Devilly, 2009) Almli et al., 2018; Dunkley et al., 2015; Y. Huang, Mohan, De Ridder, Sunaert, & Vanneste, 2018; Park et al., 2017). Using a longitudinal framework, a study recently found that childhood assaultive trauma is associated with changes in frontal alpha EEGc and subsequent AUD and PTSD symptoms, with evidence of sex specific findings in a sample of trauma-exposed and non-exposed individuals (Neale et al., 2024). Although numerous EEG studies have described differences in functional connectivity in ADRD compared to healthy participants, inconsistent results have been reported due to multiple methodological factors such as diagnostic criteria, small sample sizes and study populations limited by sex and ancestry (e.g., most of these studies have been conducted in European ancestry populations) (Fischer, Zibrandtsen, Høgh, & Musaeus, 2023). Despite these limitations, a systematic review of this literature concluded that the most consistent results involve a decrease in alpha coherence among individuals with ADRD, particularly at the left temporal region (Fischer et al., 2023). Reductions in alpha EEG coherence also appear to be one of the earliest detectable changes in individuals with ADRD (Musaeus et al., 2019). Despite the significant role *APOEε4* plays in risk for ADRD and the extensive cross-sectional research on EEGc and ADRD, no studies to date have examined the association between *APOEε4* and EEGc in adolescence longitudinally, nor concerning AUD and PTSD comorbidity.

In the current study, we examined associations between childhood trauma, *APOEε4*, and measures of brain functioning (i.e., neural connectivity) across adolescence and young adulthood (ages 12-32) stratified by sex, as well as interactions between trauma exposure and *APOEε4* on neural functioning. We also examined associations between differences in neural connectivity with downstream psychiatric (AUD and PTSD symptoms) and cognitive outcomes in young adulthood. We used data from the Collaborative Study on the Genetics of Alcoholism (COGA), a multi-site study of extended families densely affected with alcohol problems. Given our emphasis on associations between trauma, genetic risk, and neural functioning in adolescence, we focused specifically on the COGA prospective sample, comprising the adolescent and young adult offspring of the original COGA family sample. We hypothesized that childhood trauma and *APOEε4* would be associated with differences in frontal alpha EEGc, with the most robust effects observed for individuals with childhood sexual assaultive trauma, a greater dementia risk and an *APOE*ε*4 dose.* We further hypothesize that changes in neural connectivity associated with childhood trauma will associate with greater PTSD and AUD symptoms and EEG outcomes in young adulthood. We hope that findings will improve understanding of the way ADRD risk factors (genetics, alcohol use, trauma/PTSD, EEGc patterns) typically examined later in life, may manifest earlier in the lifespan. Further, findings may be used to inform novel therapies for cognitive impairments and ultimately may help to prevent the cascading risk for a wide range of health conditions related to childhood trauma.

## Method

### Participants

The Collaborative Study on the Genetics of Alcoholism (COGA) is an ongoing, multisite family study that began in 1989 and is described in detail elsewhere (Agrawal et al., 2023; Dick, Bucholz et al., 2023; Meyers, Brislin et al., 2023). From 2019-2024 COGA studied the adolescent and young adult offspring from these families in a prospective study with multidomain assessments including clinical interview, neurophysiological and neuropsychological measures, and genetic data). A total of 3,715 offspring (14,495 assessments) from families densely affected with AUD and community comparison families who had at least one parent interviewed in an earlier phase of the COGA study, were enrolled between the ages of 12-22. New participants were added as they reached the age of 12. COGA re-assessed participants ~every two years. In the present study sample, we included 837 prospective study participants (53% female) who were 1) between the ages of 12-16 at first EEG assessment, 2) endorsed exposure to at least one traumatic event by Follow-up 3, 3) had genotypic data available, and 4) had 3 or more EEG assessments. Childhood trauma was defined as trauma occurring by age 12 to ensure that traumatic exposure occurred prior to baseline EEG assessment.

### Procedure

A comprehensive battery was administered that included the adult Semi-Structured Assessment for the Genetics of Alcoholism (SSAGA) or the age-appropriate adolescent SSAGA (cSSAGA-A) and parent version (cSSAGA-P), if participants were under age 18 (Bucholz et al., 1994). The SSAGA and cSSAGA-A covered substance use problems as well as other psychiatric disorders, including PTSD. A brain function battery was administered at each assessment, which included neuropsychological tasks and neurophysiological measures during resting state (EEG coherence, EEGc) (Meyers et al., 2023). All procedures were reviewed and approved by the Institutional Review Board at each of six collection sites and informed consent was obtained at each stage of data collection.

### Measures

#### *APOEε4* genotype

Genotyping, imputation, and quality control for the COGA sample are summarized in the Supplementary materials and described in further detail in Johnson et al. (2023). *APOE* haplotypes were derived using two SNPs (rs429358 and rs7412), from which the number of ε*4* alleles were coded (0-2) and treated as a linear variable. There were 244 *APOEε4* carriers (28.14%) in the analytic sample.

#### EEG recording, data processing and measures

Procedures have been detailed previously (Meyers, Brislin et al., 2023) and are summarized in the Supplementary materials. In this study, EEG coherence (EEGc) from bipolar pairs at three frontal sites (left intra-hemispheric frontal-central (LFC: FZ-CZ--F3-C3), right intra-hemispheric frontal-central (RFC: FZ-CZ--F4-C4), and prefrontal inter-hemispheric (PFI: F8-F4--F7-F3) during eyes closed resting state were measured.

#### Childhood trauma

Lifetime history of 21 potentially traumatic events (DSM-IV Criterion A) were drawn from cSSAGA-A baseline assessments. Based on evidence that interpersonal assaultive events are more “potent” than non-assaultive events, that traumatic events cluster together, and to remain consistent with prior studies (Meyers et al., 2019; Neale et al., 2024; Subbie-Saenz de Viteri et al., 2020), we constructed three non-mutually exclusive variables representing report of 1) one or more childhood physical assaultive traumas (CPAT; stabbed, shot, mugged, threatened with a weapon, robbed, kidnapped, held captive), 2) childhood sexual assaultive traumas (CSAT; rape or molestation), and 3) childhood non-assaultive traumas (CNAT; life-threatening accident, disaster, witnessing someone seriously injured or killed, unexpectedly finding a dead body). We limited childhood trauma to those incidents reportedly experienced by age 12 to ensure that traumatic exposure occurred prior to baseline EEGc measures. CPAT, CSAT, and CNAT were endorsed by 12.22%, 5.00%, and 34.49% of the sample, respectively. Lifetime trauma exposure was based on endorsement of any of the above-described traumatic events from baseline through Follow-up 3.

#### AUD and PTSD Symptoms

Although the SSAGA or cSSAGA-A was given at all assessment timepoints, *lifetime maximum* DSM-5 AUD symptom count (AUDsx; range 0-11) and, for those who endorsed a Criterion A event, *lifetime maximum* DSM-IV PTSD Criterion B-D symptom count scores (PTSDsx; range 0-20) were assessed at each wave. In this study, we used lifetime symptom counts reported at Follow-up 3 (M_age_ = 22) to allow sufficient opportunity for AUDsx to present in late adolescence/young adulthood. The lifetime measure of PTSD and AUD severity was operationalized by lifetime maximum symptom counts, as opposed to time-point specific symptom severity.

#### Covariates

Based on literature that suggests socioeconomic status (SES) can impact brain development (Hackman, Farah, & Meaney, 2010), we covaried for family SES as indexed by parental report of highest education in COGA Phase 1-3. Analyses including *APOEε4* also covaried for the first ten genetic principal components (PCs) derived from the genome-wide data to account for population structure (Price et al., 2006).

### Data Analysis

All analyses were stratified by sex and clustered by family to account for relatedness. Primary analyses examined discrete childhood trauma variables (CPAT, CSAT, CNAT) and *APOEε4* as predictors of intercept and slope of frontal alpha EEG inter- and intra-hemispheric coherence during adolescence and young adulthood using latent growth curve models (LGCM) for those with three or more EEG assessments. LGCM models were computed using Mplus version 8.9 (Muthén & Muthén, 1998-2017), which employs full information maximum likelihood to account for missing data.

EEGc pairs at three frontal sites (left intra-hemispheric frontal-central (LFC: FZ-CZ--F3-C3), right intra-hemispheric frontal-central (RFC: FZ-CZ--F4-C4), and prefrontal inter-hemispheric (PFI: F8-F4--F7-F3) were examined in separate models. We modeled both linear and quadratic growth components for EEGc. The models that included the quadratic term did not converge; thus, we moved ahead with a linear slope term. All subsequent use of the term slope refers to linear slope.

Next, we examined the association between childhood trauma exposure, *APOEε4*, and the intercepts and slopes of EEGc. The intercept estimates individual variation in starting values which may reflect variations in genetic risk and different types of exposures prior to the measurement period, as well as other trait differences. The slope reflects how change in EEGc occurs over time, on average. Given the known co-occurrence of different types of trauma exposures, the models allowed for simultaneous testing of associations between trauma exposure types and EEGc intercept and slope. The models also evaluated whether intercept and growth parameters in EEGc were associated with AUDsx and PTSDsx. The direct effect of *APOEε4* on AUDsx and PTSDsx was also included in the models. Figure 1 displays a path diagram for the main effects model. Interactions between *APOEε4* and childhood trauma variables were examined in separate interaction effects models.

**Figure 1.**
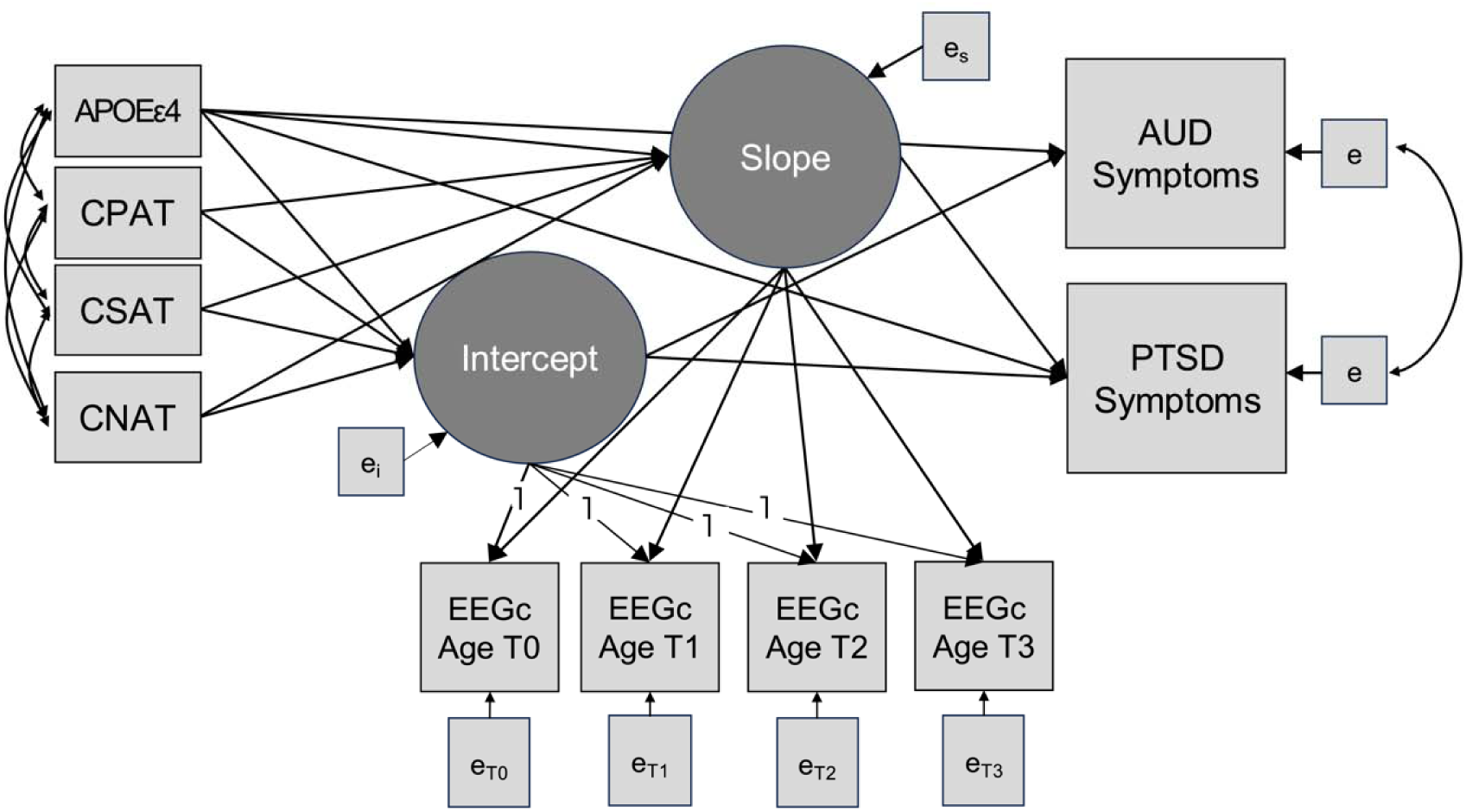
Path diagram of latent growth curve model estimating associations between childhood trauma, *APOEε4*, intercept and slope of EEG coherence, and Follow-up 3 AUD and PTSD symptoms *Note*. Growth curves were estimated based on individually varying age at the time of each EEG coherence observation. Parental education and ten ancestry principal components were included as covariates but are not depicted in the path diagram. Note, CNAT = Childhood non-assaultive trauma, CSAT = Childhood sexual assaultive trauma, CPAT = Childhood physical assaultive trauma, EEGc = EEG coherence, PTSD = Posttraumatic stress disorder, AUD = alcohol use disorder.

## Results

### Descriptive statistics

Table 1 provides sample descriptive information for demographic and key analytic variables compared by sex. The sample (N=867) was 52.94% female. There were no differences in the proportion of racial/ethnic groups between males and females in the analytic sample. Just over half the sample identified as White and about a third of the sample identified as Black/African American. Females were slightly younger than males at baseline, though the average age for both groups at first assessment was 14 years of age. Male participants reported significantly greater maximum drinks in 24 hours and AUD symptoms at follow-up 3 compared to female participants. Male participants were also more likely to endorse childhood non-assaultive and physical assaultive trauma, whereas females were more likely to report sexual assaultive trauma. Female participants also reported significantly more PTSD symptoms (mean = 1.44) than males (mean = 0.74) at Follow-up 3, though the overall number of PTSD symptoms was relatively low in both groups. Females had slightly higher *APOEε4* dose, but there were no differences in the proportion of female versus male *APOEε4* carriers.

**Table 1.** Descriptive information for sample demographic characteristics and key variables, compared by sex.

| Mean (SD) or N (%) | Male<br>Baseline<br>N=408 | Female<br>Baseline<br>N=459 | $z/t/\chi^2$ | *P* |
| --- | --- | --- | --- | --- |
| Race/ethnicity |  |  |  |  |
| White | 224 (54.90) | 254 (55.34) | 0.17 | .898 |
| Black/African American | 137 (33.58) | 166 (36.17) | 0.64 | .425 |
| Asian | 3 (0.74) | 4 (0.87) | 0.50 | .823 |
| Hispanic | 55 (13.48) | 46 (10.02) | 2.51 | .113 |
| N at follow-up 1 | 383 (93.87) | 438 (95.42) | -1.02 | .308 |
| N at follow-up 2 | 356 (87.25) | 406 (88.45) | -0.54 | .589 |
| N at follow-up 3 | 290 (71.08) | 329 (71.68) | -0.19 | .849 |
| Age at baseline | 14.35 (1.56) | 14.03 (1.56) | 2.97 | **.003** |
| Age at follow-up 1 | 17.65 (2.65) | 17.41 (2.54) | 1.31 | .190 |
| Age at follow-up 2 | 20.50 (2.81) | 20.45 (3.10) | 0.22 | .825 |
| Age at follow-up 3 | 22.64 (3.06) | 22.40 (3.42) | 0.93 | .351 |
| Parental education (years) | 13.07 (2.22) | 12.98 (2.12) | 0.56 | .580 |
| Ever drink | 383 (93.87) | 431 (93.90) | -0.02 | .984 |
| Maximum drinks | 19.29 (16.22) | 12.16 (11.62) | 7.36 | **<.001** |
| Initiated regular drinking | 338 (82.84) | 364 (79.30) | 1.76 | .185 |
| Onset age of regular drinking | 17.74 (2.46) | 18.10 (2.38) | -1.95 | .052 |
| AUD symptoms at follow-up 3 | 1.20 (3.57) | 0.73 (1.49) | 3.83 | **<.001** |
| Non-assaultive trauma | 154 (37.75) | 145 (31.59) | 5.44 | **.020** |
| Physical-assaultive trauma | 73 (17.89) | 33 (7.19) | 25.30 | **<.001** |
| Sexual-assaultive trauma | 7 (1.72) | 36 (7.84) | 16.26 | **<.001** |
| PTSD symptoms at follow-up 3 | 0.74 (2.58) | 1.44 (3.63) | -2.84 | **.005** |
| APOEε4 dose | 0.29 (0.49) | 0.36 (0.57) | -1.97 | **.050** |
| APOEε4 carrier | 105 (25.74) | 139 (30.28) | 1.78 | .183 |
| *Note:* AUD = alcohol use disorder, and PTSD = post-traumatic stress disorder. Maximum drinks, AUD symptoms, PTSD symptoms were lifetime measures assessed at follow-up 3. APOEε4 dose is the mean number of APOEε4 alleles. APOEε4 carrier represents the number of participants who were carriers of at least one APOEε4 allele. **Bold** text indicates p-value less than .05. **Bold italic** text indicates p-value less than .01. |  |  |  |  |

### Childhood trauma and EEG connectivity trajectories

Results of the LGCM estimating the unconditional mean intercept and linear slope of EEGc are provided in Supplementary Table 1. There was a significant positive mean intercept and slope across all three EEGc pairs, suggesting that on average, EEGc tends to increase across adolescence. Results of LGCM demonstrating main effect associations between childhood trauma and EEGc are provided in Table 2. In females, we found that CSAT was positively associated with intercept (ß= 0.10 to 0.13, *p* < 0.001 to 0.002) and negatively associated with slope (ß= −0.05 to −0.07, *p* < 0.001 to 0.005) of all three EEGc pairs. This suggests that exposure to CSAT was associated with higher initial values of frontal EEGc, but diminished growth in EEGc across development. CNAT was negatively associated with intercept of LFC (ß= −0.05, *p* = 0.001) and RFC (ß= −0.04, *p* = 0.015) EEGc, suggesting lower LFC and RFC initial EEGc values in females who endorsed CNAT. In females, CPAT was positively associated with intercept of PFI EEGc (ß= 0.07, *p* = 0.007), suggesting those CPAT was associated with higher baseline PFI neural connectivity in females. In males, CPAT was negatively associated with intercept of LFC EEGc (ß= −0.17, *p* = 0.037), but positively associated with slope of LFC EEGc (ß= 0.07, *p* = 0.007). CNAT was negatively associated with intercept of RFC EEGc (ß= −0.04, *p* = 0.046) in males. No significant associations between CSAT and EEGc were observed in males.

**Table 2.** Linear Growth Model Results for the Effect of Childhood Trauma Exposure and APOEε4 on Slope and Intercept of Three Frontal Alpha EEG Coherence Pairs and Subsequent AUD and PTSD symptoms in Trauma-Exposed Adolescent Male and Female COGA Participants.

|  | Left frontal-central (LFC) EEG coherence<br>FZ-CZ--F3-C3 |  |  |  | Right frontal-central (RFC) EEG coherence<br>FZ-CZ--F4-C4 |  |  |  | Prefrontal interhemispheric (PFI) EEG coherence<br>F8-F4--F7-F3 |  |  |  |
| --- | --- | --- | --- | --- | --- | --- | --- | --- | --- | --- | --- | --- |
|  | Males |  | Females |  | Males |  | Females |  | Males |  | Females |  |
|  | Beta (S.E.) | *p* | Beta (S.E.) | *p* | Beta (S.E.) | *p* | Beta (S.E.) | *p* | Beta (S.E.) | *p* | Beta (S.E.) | *p* |
| Associations with Intercept and Slope of Alpha EEG Coherence |  |  |  |  |  |  |  |  |  |  |  |  |
| CNAT on Intercept | -0.05 (0.04) | .198 | -0.05 (0.01) | .001 | -0.04 (0.02) | .045 | -0.04 (0.02) | .015 | -0.04 (0.14) | .785 | -0.02 (0.01) | .116 |
| CSAT on Intercept | 0.24 (0.22) | .282 | 0.10 (0.03) | .002 | 0.03 (0.14) | .844 | 0.12 (0.04) | .002 | 0.048 (0.13) | .701 | 0.13 (0.03) | <.001 |
| CPAT on Intercept | -0.17 (0.08) | .037 | 0.04 (0.03) | .109 | 0.00 (0.03) | .884 | 0.04 (0.02) | .111 | 0.01 (0.08) | .902 | 0.07 (0.03) | .007 |
| *APOEε4* on Intercept | -0.11 (0.04) | .003 | -0.07 (0.04) | .066 | -0.05 (0.04) | .216 | -0.08 (0.04) | .024 | -0.07 (0.32) | .827 | 0.06 (0.05) | .207 |
| Parental education on Intercept | 0.00 (0.01) | .844 | -0.00 (0.00) | .674 | 0.01 (0.01) | .582 | -0.00 (0.00) | .397 | 0.00 (0.02) | .875 | -0.00 (0.00) | .201 |
| CNAT on Slope | 0.03 (0.02) | .238 | 0.00 (0.01) | .891 | 0.02 (0.01) | .127 | 0.00 (0.01) | .749 | 0.02 (0.08) | .795 | -0.01 (0.01) | .456 |
| CSAT on Slope | -0.10 (0.12) | .424 | -0.05 (0.02) | .002 | 0.00 (0.08) | .991 | -0.05 (0.02) | .005 | -0.01 (0.07) | .870 | -0.07 (0.02) | <.001 |
| CPAT on Slope | 0.11 (0.05) | .021 | -0.02 (0.01) | .135 | 0.01 (0.02) | .477 | -0.02 (0.01) | .135 | -0.01 (0.05) | .868 | -0.01 (0.02) | .551 |
| *APOEε4* on Slope | 0.06 (0.02) | .002 | 0.03 (0.02) | .081 | 0.02 (0.02) | .223 | 0.04 (0.02) | .021 | 0.04 (0.18) | .823 | -0.02 (0.03) | .410 |
| Parental education on Slope | -0.00 (0.01) | .650 | 0.00 (0.00) | .105 | -0.00 (0.01) | .605 | 0.00 (0.00) | .108 | -0.00 (0.01) | .525 | 0.00 (0.00) | .257 |
| Associations with AUD and PTSD Symptoms |  |  |  |  |  |  |  |  |  |  |  |  |
| Intercept on AUD Symptoms | -0.54 (0.32) | .090 | -0.58 (0.52) | .265 | -1.97 (0.34) | <.001 | -0.82 (0.52) | .115 | -1.64 (9.71) | .866 | 0.457 (0.513) | .373 |
| Slope on AUD Symptoms | -0.13 (0.28) | .641 | -3.51 (1.25) | .005 | 0.29 (1.16) | .805 | -4.21 (1.29) | .001 | 2.14 (1.06) | .044 | -1.028 (0.728) | .158 |
| Intercept on PTSD Symptoms | 0.09 (0.07) | .183 | 0.27 (0.12) | .026 | 0.23 (0.09) | .009 | 0.29 (0.13) | .024 | 0.15 (3.88) | .969 | 1.224 (0.101) | <.001 |
| Slope on PTSD Symptoms | -0.07 (0.07) | .337 | -4.74 (1.17) | <.001 | -0.43 (0.34) | .213 | -4.83 (1.32) | <.001 | -0.71 (0.63) | .262 | -1.799 (0.472) | <.001 |
| *APOEε4* on AUD Symptoms | -0.06 (0.08) | .435 | 0.14 (0.11) | .189 | -0.09 (0.12) | .421 | 0.18 (0.13) | .149 | 0.00 (0.08) | .968 | -0.20 (1.63) | .902 |
| *APOEε4* on PTSD Symptoms | -0.00 (0.02) | .930 | 0.18 (0.13) | .172 | 0.00 (0.03) | .927 | 0.23 (0.14) | .094 | -0.00 (0.01) | .801 | 0.02 (0.46) | .963 |
| Observations | 408 | - | 459 | - | 408 | - | 459 | - | 408 | - | 459 | - |
| AIC | 3387.67 | - | 4524.20 | - | 3435.75 | - | 4523.34 | - | 3317.11 | - | 4133.07 | - |
| BIC | 3669.45 | - | 4815.32 | - | 3717.53 | - | 4814.46 | - | 3598.89 | - | 4424.20 | - |
| H<sub>0</sub> log-likelihood | -1622.84 | - | -2191.10 | - | -1646.87 | - | -2190.67 | - | -1587.55 | - | -1995.54 | - |
| H<sub>1</sub> maximum log-likelihood | 10803.47 | - | 20941.41 | - | 10775.14 | - | 12047.75 | - | 10848.44 | - | 12253.71 | - |

Note: CNAT = Childhood non-assaultive trauma, CSAT = Childhood sexual assaultive trauma, CPAT = Childhood physical assaultive trauma. AIC= Akaike Information Criterion, BIC=Bayesian Information Criterion. **Bold** text indicates p-value less than .05. ***Bold italic*** text indicates p-value less than .01.

### *APOEε4* main effects and interactions between *APOE***ε***4* and childhood trauma on EEG connectivity trajectories

Associations between *APOEε4* and EEGc intercept and slope are displayed in Table 2. In females, we found that *APOEε4* was negatively associated with RFC EEGc intercept (ß= −0.08, *p* = 0.024) and positively associated with RFC EEGc slope (ß= 0.04, *p* = 0.021). In males, *APOEε4* was significantly negatively associated with intercept (ß= −0.10, *p* = 0.005) and positively associated with slope of LFC EEGc (ß= 0.06, *p* = 0.004). We also examined interactions between childhood trauma variables and *APOEε4* on intercept and slope of EEGc, which are displayed in Supplementary Table 2. Although some interaction effects were nominally significant, these associations were no longer significant after correcting for multiple testing (Benjamini & Hochberg, 1995).

### Associations between EEG trajectories and young adult PTSD and AUD symptoms

Associations between EEGc intercept and slope with PTSD and AUD symptoms at Follow-up 3 are displayed in Table 2. In females, intercept of LFC (ß= 0.27, *p* = 0.026) and RFC (ß= 0.29, *p* = 0.024) EEGc were positively associated with PTSD symptoms. Slope of LFC, RFC, and PFI EEGc were negatively associated with PTSD symptoms (ß= −4.23 to −1.80, *p* < 0.001). Slope of LFC (ß= −3.51, *p* = 0.005) and RFC (ß= −4.21, *p* = 0.001) EEGc were also negatively associated with AUD symptoms in females. In males, intercept of RFC EEGc was negatively associated AUD symptoms (ß= −1.97, *p* < 0.001), but positively associated with PTSD symptoms (ß= 0.23, *p* = 0.009). We found no direct associations between *APOEε4* and PTSD or AUD symptoms.

## Discussion

This study is one of the first to examine associations between childhood trauma, Alzheimer’s genetic risk and *APOEε4* dose with frontal EEG coherence (EEGc) in adolescence, as well as their joint association with PTSD and AUD in young adulthood. Results of the association between childhood trauma and EEGc were largely consistent with what was found in a related analysis (Neale et al., 2024), wherein childhood sexual assaultive trauma in females and childhood physical assaultive trauma in males were linked to differences in frontal alpha EEGc. Importantly, the present analyses also accounted for one of the largest genetic risk factors for Alzheimer’s disease, *APOEε4*, which is rarely examined in adolescent neurodevelopment despite evidence that cognitive differences related to *APOEε4* (Chang et al., 2016; Iacono & Feltis, 2019) as well as trauma and alcohol use during adolescence (Carrion & Wong, 2012; Herringa, 2017; Lees, Meredith, Kirkland, Bryant, & Squeglia, 2020). We found that in both sexes, *APOEε4* was associated with lower baseline EEGc and increased slope of EEGc, though these effects varied by brain region in males and females. Moreover, there were strong links between EEGc patterns and symptoms of PTSD and AUD in females, but in males EEGc had more pronounced associations with AUD symptoms.

### Childhood trauma and neurodevelopment

Consistent with our hypotheses, childhood trauma prior to age 13 was associated with differences in neural connectivity, supporting theoretical and empirical research that traumatic events occurring early in life may alter neurodevelopment (Bremner, 2006; Cook et al., 2009). In females, we found that sexual trauma was associated with higher baseline and decreased growth in all three EEGc pairs. In males, we found lower baseline but increased growth of LFC EEGc in those with childhood physical assaultive trauma. Interestingly, non-assaultive trauma was also associated with decreased baseline LFC EEGc in males and females, as well as decreased RFC EEGc baseline in females only. These effects suggest that different types of trauma exposure may have differential effects on neural connectivity, wherein sexual assaultive trauma in females is associated with heightened baseline EEGc and physical assaultive trauma in males and non-assaultive trauma in both sexes are associated with lower baseline EEGc. This aligns with literature that indicates threat versus deprivation may contribute to different types of alterations in neurodevelopment (McLaughlin, Sheridan, & Lambert, 2014), as well as literature that the impact of trauma on neurodevelopment may vary by sex (Bremner, 2006; Helpman et al., 2017). Indeed, these relationships may also be influenced by sex differences in the prevalence of trauma exposure type, as well as the increased risk of PTSD following trauma exposure in females (Kessler, Davis, & Kendler, 1997). Thus, while childhood trauma clearly impacts brain functioning, the nature of these associations is highly complex and dependent on a number of factors including type of trauma exposure, sex, and other factors unaccounted for in this study, such as timing, frequency, and intensity of traumatic events (Helpman et al., 2017).

### APOEε4

Our results also indicate that *APOEε4* is associated with neural changes in youth, paralleling findings observed in older individuals with AD. We found that *APOEε4* was associated with decreased intercept of LFC alpha EEG coherence in males and decreased RFC alpha EEG coherence in females. A systematic review of studies of EEG coherence in Alzheimer’s disease (Fischer et al., 2023) found strong evidence of decreased alpha EEG coherence among individuals with AD compared to controls, with effects most pronounced in left frontal regions. There is also evidence that *APOEε4* carriers have decreased alpha EEGc in frontal regions (Jelic et al., 1997), as well as changes in other EEG measures, such as more pronounced decreases in alpha EEG power (Ponomareva, Korovaitseva, & Rogaev, 2008) and increased EEG complexity (Gutiérrez-de Pablo et al., 2020). However, existing studies of *APOEε4* and EEG phenotypes have typically used smaller (N<100) cross-sectional studies in older adults. Thus, our finding that *APOEε4* was associated with decreased intercept but increased slope in adolescence provides a novel contribution to our understand of the way these associations may change over time. Further longitudinal research is needed to determine whether decreased alpha EEG coherence is associated with neuropsychological, and structural and functional brain changes, as well as whether the phenotype may serve as an indicator of individuals who may benefit from early cognitive interventions.

### PTSD and AUD Outcomes

In females, CSAT was linked to decreased growth of EEGc, which was subsequently linked to greater PTSD and AUD symptoms in young adulthood. This suggests that the changes associated with early childhood sexual trauma in women are associated with differences in neural connectivity that may relate to poorer psychiatric functioning in young adulthood. This finding is consistent with literature suggesting childhood trauma, particularly sexual trauma, can leave lasting signatures on neurodevelopment (Edwards, 2018). Although the ability to test causal associations is limited in this observational study, the associations do point to a possibility that neural connectivity may be a biomarker linking childhood trauma to later mental health outcomes. Interestingly, the nature of these associations differs between men and women. In males, only baseline values of RFC EEGc were significantly associated with PTSD and AUD symptoms in young adulthood. Following trauma exposure, women are more likely to develop PTSD and comorbid PTSD and AUD than men (Kessler et al., 1997; Peltier et al., 2022), though the sex-differences may also be in part due to the higher prevalence of childhood physical compared to sexual assaultive trauma in males (Erol & Karpyak, 2015; Helpman et al., 2017).

### Strengths, Limitations, and Future Directions

While this is one the first studies to use a longitudinal design to model EEGc trajectories during adolescence, addressing a gap in static, cross-sectional research examining *APOEε4*-by-trauma interactions in a diverse cohort, these findings should be considered within the context of several study constraints. First, in regard to the sample and participant selection—this study is limited to trauma exposed participants, many of whom are from AUD-affected families, which may limit generalizability. Next, retrospective trauma reports could introduce recall bias and some aspects of childhood trauma, specifically deprivation, are not well defined in this sample. Finally, interaction effects between trauma and *APOEε4* did not withstand a multiple-test-correction, suggesting potential limitations in statistical power and/or other unexplored mediators/moderators (e.g., resilience factors, social support, alcohol initiation/use) that should be explored in future analyses. Relatedly, in addition to EEG coherence, future studies should incorporate other measures of neural dynamics, including EEG-based and fMRI-based measures, in addition to neuropsychological assessments of cognitive function.

In summary, the present study aimed to understand associations between childhood trauma, genetic risk for Alzheimer’s disease via *APOEε4*, adolescent neural connectivity, and young adult AUD and PTSD. We found that there are sex- and trauma-specific associations between childhood trauma, *APOE*ε*4,* and frontal alpha EEG coherence, and that those associations are also linked to differences in AUD and PTSD symptoms in young adulthood. These findings may be important for identifying individuals at greater risk for cognitive and psychiatric harms related to early childhood trauma and may also be used to inform neurophysiological treatments (e.g., transcranial magnetic stimulation) to reduce symptoms associated with traumatic stress. Future research that investigates mediating mechanisms (e.g., inflammation, epigenetics, alcohol initiation) and other genetic risk factors (e.g., polygenic scores for AD) would further expand understanding of links between trauma, genetics, neural connectivity and long-term psychiatric and brain health outcomes. Furthermore, studies that include richer phenotyping of trauma exposure, other influential adverse childhood experiences, and social determinants of health would better inform the degree to which trauma is associated with normative and atypical aging across the lifespan. Overall, our study increases understanding of the complex associations between childhood trauma, genetics, and brain functioning during the important, yet understudied period of adolescent neurodevelopment, and draws links to psychiatric functioning later in life. Findings may be used to improve clinical understanding of sex differences in the relationship between trauma and *APOEε4* and neurophysiology, as well as potentially inform intervention and treatment of childhood trauma to mitigate long-term impacts on mental health and aging.

## Data Availability

All data produced in the present study are available upon reasonable request to the authors

## Acknowledgements

The Collaborative Study on the Genetics of Alcoholism (COGA), Principal Investigators B. Porjesz, V. Hesselbrock, T. Foroud; Scientific Director, A. Agrawal; Translational Director, D. Dick, includes ten different centers: University of Connecticut (V. Hesselbrock); Indiana University (H.J. Edenberg, T. Foroud, Y. Liu, M.H. Plawecki); University of Iowa Carver College of Medicine (S. Kuperman, J. Kramer); SUNY Downstate Health Sciences University (B. Porjesz, J. Meyers, C. Kamarajan, A. Pandey); Washington University in St. Louis (L. Bierut, J. Rice, K. Bucholz, A. Agrawal); University of California at San Diego (M. Schuckit); Rutgers University (J. Tischfield, D. Dick, R. Hart, J. Salvatore); The Children’s Hospital of Philadelphia, University of Pennsylvania (L. Almasy); Icahn School of Medicine at Mount Sinai (A. Goate, P. Slesinger); and Howard University (D. Scott). Other COGA collaborators include: L. Bauer (University of Connecticut); J. Nurnberger Jr., L. Wetherill, X., Xuei, D. Lai, S. O’Connor, (Indiana University); G. Chan (University of Iowa; University of Connecticut); D.B. Chorlian, J. Zhang, P. Barr, S. Kinreich, Z. Neale, G. Pandey (SUNY Downstate); N. Mullins (Icahn School of Medicine at Mount Sinai); A. Anokhin, S. Hartz, E. Johnson, V. McCutcheon, S. Saccone (Washington University); J. Moore, F. Aliev, Z. Pang, S. Kuo (Rutgers University); A. Merikangas (The Children’s Hospital of Philadelphia and University of Pennsylvania); H. Chin and A. Parsian are the NIAAA Staff Collaborators. We continue to be inspired by our memories of Henri Begleiter and Theodore Reich, founding PI and Co-PI of COGA, and also owe a debt of gratitude to other past organizers of COGA, including Ting-Kai Li, P. Michael Conneally, Raymond Crowe, and Wendy Reich, for their critical contributions. This national collaborative study is supported by NIH Grant U10AA008401 from the National Institute on Alcohol Abuse and Alcoholism (NIAAA) and the National Institute on Drug Abuse (NIDA).

