## Supplementary Materials for "Childhood Trauma and *APOEε4* are Associated with Adolescent Brain Function, Posttraumatic Stress, and Alcohol-related Outcomes"

*EEG Recording and Data Processing.* Briefly, resting (eyes-closed) EEG was recorded for 4.25 min; a continuous interval of 256 seconds was analyzed. Each subject wore a fitted electrode cap containing a 61-channel montage of scalp electrodes (Electro-Cap International Inc). The nose served as reference and the ground electrode was placed on the forehead. Eye movements are monitored with electrodes above and below left eye. EEG was recorded with the subjects seated comfortably in a dimly lit sound-attenuated temperature-regulated booth. They were instructed to keep their eyes closed and remain relaxed, but not to fall asleep. Electrode impedances were maintained below 5 kΩ. EEG activity was amplified by a factor of 10,000 on Neuroscan amplifiers (Synamps2), filtered between 0.02 Hz and 100 Hz and sampled at 500 Hz using the Neuroscan software system running on 186 PCs. Identical procedures were performed at all collection sites at baseline and each follow-up assessment. Digitized electrophysiological data underwent quality control and were edited for known artifacts (movement, EMG, eye movement, DC shifts); filtering and ocular correction were implemented where required. Between 19-64 channels according to the 10-20 International system were used for analysis. Bipolar electrode pairs were derived to reduce volume conduction effects. Conventional Fourier transform methods were used to calculate EEG interhemispheric and intrahemispheric coherence at 27 bipolar pairs in the following frequency bands: theta (3-7 Hz), alpha (8-12 Hz), and beta (13-20 Hz). Additional information on EEG methods are available (Meyers, Brislin et al., 2023). Supplementary Figure 1 depicts the three bipolar electrode pairs examined in the present study.

**Supplementary Figure 1.**

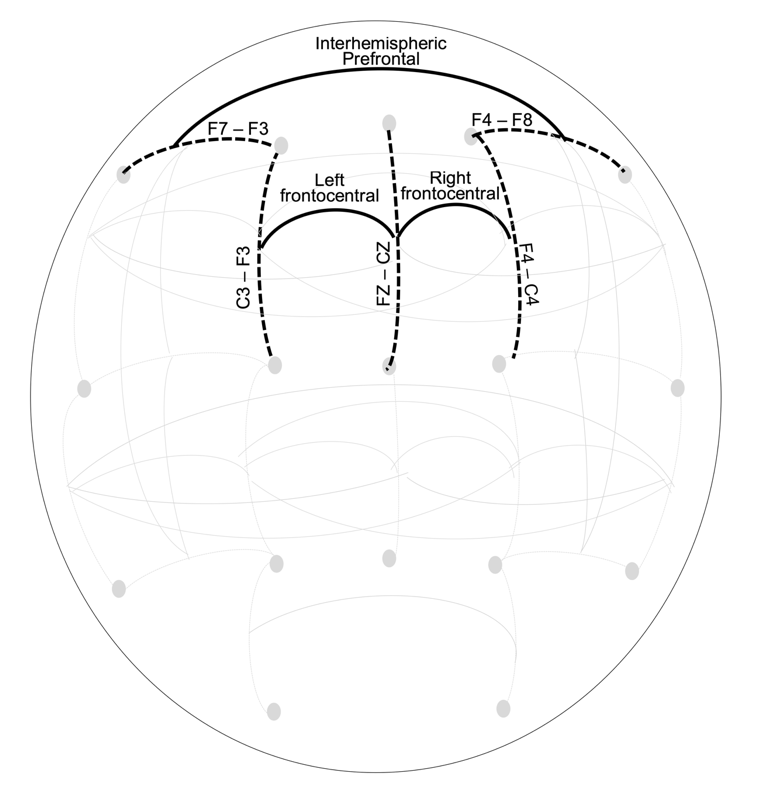

*Note*. This figure displays a schematic of bipolar electrode pairs (indicated by black dotted lines) and coherence pairs (indicated by black solid lines) derived between bipolar electrode pairs. This figure was previously published in *Psychological Medicine*.

*Genotyping and quality control procedures.* Briefly, four different arrays were used to genotype COGA samples, with some overlap to allow for quality control. SNPs with a genotyping rate <98%, Hardy-Weinberg equilibrium violations (p<10^-6^), or with minor allele frequency (MAF) less than 3% were excluded from analyses. Mendelian inconsistencies were removed (O’Connell & Weeks, 1998), after which data were imputed to 1000 genomes (EUR and AFR, Phase 3, b37, October 2014; build hg19) using SHAPEIT (Delaneau, Zagury, & Marchini, 2013) and IMPUTE2 (van Leeuwen et al., 2015). Following imputation, genotype probabilities ≥ 0.90 were changed to genotypes. Mendelian errors in the imputed SNPs were reviewed and resolved. SNPs with an imputation information score < 0.30 or MAF < 0.03 were excluded from subsequent analysis.

| **Supplementary Table 1.** Results of the Unconditional Linear Growth Model for Repeated Measures of Three Frontal Alpha EEG Coherence Pairs in Trauma-Exposed Adolescent Male and Female COGA Participants. | | | | | | | | | | | | | |
| --- | --- | --- | --- | --- | --- | --- | --- | --- | --- | --- | --- | --- | --- |
|  | Left frontal-central (LFC) EEG coherence FZ-CZ--F3-C3 | | | | Right frontal-central (RFC) EEG coherence  FZ-CZ--F4-C4 | | | | | Prefrontal interhemispheric (PFI) EEG coherence  F8-F4--F7-F3 | | | |
|  | Males | | Females | | Males | | Females | | | Males | | Females | |
|  | Beta (S.E.) | *p* | Beta (S.E.) | *p* | Beta (S.E.) | *p* | Beta (S.E.) | *p* | Beta (S.E.) | | *p* | Beta (S.E.) | *p* |
| **Unconditional Linear Growth Model** | | | | | | | | | | | | | |
| Intercept Mean | 0.42 (0.02) | ***<.001*** | 0.47 (0.02) | ***<.001*** | 0.44 (0.02) | ***<.001*** | 0.46 (0.02) | ***<.001*** | 0.22 (0.02) | | ***<.001*** | 0.25 (0.02) | ***<.001*** |
| Slope Mean | 0.08 (0.01) | ***<.001*** | 0.06 (0.00) | ***<.001*** | 0.07 (0.01) | ***<.001*** | 0.01 (0.00) | ***<.001*** | 0.03 (0.01) | | ***.003*** | 0.03 (0.01) | **.011** |
| Intercept Variance | 0.03 (0.02) | **.042** | 0.00 (0.02) | .911 | 0.01 (0.01) | .293 | 0.03 (0.01) | **.037** | 0.01 (0.01) | | .329 | 0.03 (0.01) | **.023** |
| Slope Variance | 0.01 (0.01) | **.037** | 0.00 (0.00) | .762 | 0.01 (0.01) | .184 | 0.00 (0.00) | .071 | 0.00 (0.00) | | .241 | 0.01 (0.00) | **.028** |
| Intercept-Slope Correlation | -0.02 (0.01) | .075 | 0.00 (0.0) | .929 | -0.01 (0.01) | .442 | -0.00 (0.00) | .081 | -0.00 (0.01) | | .635 | -0.01 (0.01) | .086 |
| Observations | 408 | - | 459 | - | 408 | - | 459 | - | 408 | | - | 459 | - |
| AIC | -1264.87 | - | -788.00 | - | -1241.78 | - | -809.73 | - | -1402.49 | | - | -1249.18 | - |
| BIC | -1228.76 | - | -750.84 | - | -1205.68 | - | -772.56 | - | -1366.39 | | - | -1212.02 | - |
| H_0_ log-likelihood | 641.43 | - | 403.00 | - | 629.89 | - | 413.86 | - | 710.25 | | - | 633.59 | - |
| H_1_ maximum log-likelihood | 647.90 | - | 405.77 | - | 639.13 | - | 409.65 | - | 423.43 | | - | 639.39 | - |
| Note: **Bold** text indicates p-value less than .05. ***Bold italic*** text indicates p-value less than .01. | | | | | | | | | | | | | |

| **Supplementary Table 2.** Interactions Between Childhood Trauma and APOEε4 on Intercept and Slope of EEG Coherence | | | | | | |
| --- | --- | --- | --- | --- | --- | --- |
|  | Males | | | Females | | |
|  | Beta (S.E.) | *p* | Adjusted *p* | Beta (S.E.) | *p* | Adjusted *p* |
| Intercept of Left Frontal-central EEGc (FZ-CZ--F3-C3) | | | | | | |
| CNATx *APOEε4* | 0.07 (0.07) | .310 | .370 | -0.04 (0.03) | .115 | .314 |
| CPATx *APOEε4* | -0.07 (0.14) | .621 | .621 | -0.11 (0.08) | .164 | .249 |
| CSATx *APOEε4* | 0.68 (0.45) | .135 | .390 | 0.02 (0.06) | .799 | .799 |
| Slope of Left Frontal-central EEGc (FZ-CZ--F3-C3) | | | | | | |
| CNATx *APOEε4* | -0.05 (0.04) | .187 | .739 | 0.01 (0.01) | .493 | .739 |
| CPATx *APOEε4* | 0.05 (0.09) | .590 | .590 | 0.08 (0.04) | **.034** | .156 |
| CSATx *APOEε4* | -0.36 (0.24) | .136 | .272 | -0.03 (0.04) | .426 | .511 |
| Intercept of Right Frontal-central EEGc (FZ-CZ--F4-C4) | | | | | | |
| CNATx *APOEε4* | 0.05 (0.05) | .370 | .370 | -0.03 (0.02) | .157 | .314 |
| CPATx *APOEε4* | -0.11 (0.08) | .158 | .249 | -0.09 (0.07) | .166 | .249 |
| CSATx *APOEε4* | 0.65 (0.39) | .100 | .390 | 0.06 (0.06) | .301 | .452 |
| Slope of Right frontal-central EEGc (FZ-CZ--F4-C4) | | | | | | |
| CNATx *APOEε4* | -0.03 (0.03) | .314 | .739 | 0.01 (0.01) | .565 | .739 |
| CPATx *APOEε4* | 0.07 (0.05) | .125 | .250 | 0.06 (0.03) | .052 | .156 |
| CSATx *APOEε4* | -0.34 (0.22) | .113 | .272 | -0.02 (0.03) | .525 | .525 |
| Intercept of Prefrontal Interhemispheric EEGc (F8-F4--F7-F3) | | | | | | |
| CNATx *APOEε4* | 0.03 (0.03) | .355 | .370 | -0.03 (0.02) | .135 | .314 |
| CPATx *APOEε4* | -0.04 (0.04) | .252 | .302 | -0.13 (0.06) | **.036** | .216 |
| CSATx *APOEε4* | 0.20 (0.15) | .195 | .390 | 0.02 (0.06) | .737 | .799 |
| Slope of Prefrontal Interhemispheric EEGc (F8-F4--F7-F3) | | | | | | |
| CNATx *APOEε4* | -0.01 (0.02) | .739 | .739 | 0.00 (0.01) | .659 | .739 |
| CPATx *APOEε4* | 0.04 (0.03) | .172 | .258 | 0.05 (0.04) | .269 | .323 |
| CSATx *APOEε4* | -0.16 (0.08) | **.040** | .240 | -0.02 (0.03) | .387 | .511 |
| Note: CNAT = Childhood non-assaultive trauma, CSAT = Childhood sexual assaultive trauma, CPAT = Childhood physical assaultive trauma, EEGc = EEG Coherence. **Bold** text indicates p-value less than .05. ***Bold italic*** text indicates p-value less than .01. | | | | | | |
